# Qualitative assessment of the citizen science approach to foster public partnerships for pandemic preparedness and response in South and Southeast Asian countries

**DOI:** 10.1101/2024.08.21.24312377

**Authors:** Dinesh Kumar, Ingo Hauter, Felipe C. Canlas, Firli Yogiteten Sunaryoko, Gyanu Raja Maharjan, Md. Mazharul Anowar, Harjyot Khosa, Yi-Roe Tan, Peiling Yap

## Abstract

Citizen science (CS) promotes the inclusion of diverse stakeholders and offers a scientific in-depth understanding of community engagement to build trust, increase knowledge, and facilitate policymaking. Study aimed to understand concepts, practices, approaches, and sustainability issues of CS among citizens in five South and Southeast Asian countries. Qualitative study from October 2022 to March 2023 was carried in Nepal, Bangladesh, India, Philippines, and Indonesia. In each country, four focus group discussions were conducted with an overall total of 130 participants. Content analysis and coding were carried out for narrative responses of participants. Across all countries, the participants collectively comprehended the term "research" while referring to CS. Participants also related social responsibility and capacity building of citizens to CS. In terms of their contributions to pandemic response, participants stated compliance with government guidelines, helping to create awareness, and providing necessary support and assistance. Participants value personal achievement, satisfaction, happiness, and a chance to build social capital while participating in CS activities. Participants were ready to actively contribute to CS activities and share their opinions with stakeholders such as policymakers and researchers but felt that a lack of personal confidence, ineffective communication, and insufficient translation of their opinions to actions could deter them. Creation of an organization or network, provision of budget for activities, incentives to participants, and transportation assistance were considered as resources needed for the sustainability of CS. Participants expressed their readiness for CS activities considering personal and social factors, while systemic support is needed for sustained participation.

## Background

Globally, the COVID-19 pandemic has negatively affected the daily lives of people. In addition to its impact on human health, it has disrupted livelihoods and caused intense and detrimental effects on trade and economies. In resource-limited countries, especially those in the South and Southeast Asian regions, the pandemic had an overwhelming effect due to high population density, limited healthcare facilities, poor socio-economic environment, and inaccessible or inequitable social protection schemes.^1^ These countries witnessed how pandemic has threatened the progress of Sustainable Development Goals.^2^ Administrations and health systems were inundated by the pandemic as citizens, health care professionals, program managers, and policy makers battle to contain COVID-19. People’s cooperation and trust became necessary to manage, and proved challenging with the differential experiences of population groups. Adoption of various mechanisms of community engagement was thus encouraged to facilitate the implementation and adoption of COVID-19 pandemic control strategies.^3^ With a need to engage communities, risk communication was observed to be inadequate as it followed a top-down approach. Analysis observed negative impacts on people with poor experiences.^4^

The prodigious nature of COVID-19 left with immense experiences among people, health professionals, and program managers. Systematic capture and analysis of these familiarities became a rational basis for building effective community engagement to prepare and manage public health emergencies. Citizen science (CS) is a scientific approach to effectively engage people to manage public health issues. We define CS as a practice of public participation, collaboration, and co-creation in all aspects of scientific research to build trust, increase knowledge, and facilitate policymaking to address public health challenges.^5^ This approach promotes the inclusion of diverse stakeholders like local communities, researchers, policymakers, development partners, and program managers in various mechanisms of engagement with science.^6^ Due to its adoption across different domains, typological analysis of the literature has shown significant variation in the understanding and interpretation of CS as well as its multiple forms of implementation. Consequently, further studies to understand the various interpretations of CS have been proposed.^7^

CS has piqued the interest of policymakers, program managers, and researchers as a way to promote productivity and democratize the process of scientific knowledge-making, allowing collaborations between these stakeholder groups to address societal problems.^8^ CS works to improve understanding of health issues from people’s perspective and concurrently allows people to understand issues more logically and scientifically. Indeed, citizens have the right to raise and investigate their questions in the field of public health research. They also have the eagerness and capability to design, implement, and manage research projects but knowledge-sharing gap still exists between citizens and researchers.^9^ Therefore, in the assessment of the CS approach and definition, it is also important to gather perspectives of citizens to ensure resonance of such an approach with them.

As COVID-19 has had a profound impact on people’s lives and economies, a scientific approach to understand citizen’s engagement is proposed for pandemic preparedness and response. COVID-19 pandemic was used as a socially relevant use case where citizens exhibited diverse methods of participation in the management of the pandemic. Their experiences were assessed with scope and potential usability of CS approach for effective engagement in pandemic preparedness and response. Present study was carried out to understand concepts, practices, approaches, and sustainability issues of CS among citizens in five South and Southeast Asian countries. The objectives of the study were to (i) assess awareness, relatability, and acceptability of local communities to CS participatory approaches for pandemic preparedness and response; (ii) assess the level of readiness in communities to participate in CS activities; and (iii) identify barriers and facilitators for communities to participate in CS.

## Material and Methods

A sequential mixed-method study was carried out in Nepal (Rural Development Foundation), Bangladesh (Rural Development Agency), India (Health Applications), Philippines (Wireless Access to Health), and Indonesia (Climate Institute) from October 2022 to March 2023. The procedures and findings from the quantitative segment were published separately.^10^ This paper focus only on the qualitative segment based on participants’ experiences during COVID-19 pandemic. In each country, 4 focus group discussions (FGDs) were conducted with 6 to 8 participants per FGD. Number of required FGDs was based on substantiated number for effective discussion and thematic saturation. For comprehensive assessment, participants were purposively sampled to include youths (≥18 years of age), marginalized and indigenous communities (people living with HIV/AIDS, tuberculosis, malaria; ethically/socio-economic marginalized), community health workers (last mile health workers as a direct link between people and local health services), and general population. The participants were sampled from a pool of people who participated in a survey and gave consent to be re-contacted for FGD (57.2% of 2912 survey participants), as part of this mixed-method study ^10^. Participants less than 18 years of age, with severe mental health conditions, or not conversant in English or the local languages were excluded.

### Data collection tools and technique

Project staff carrying out the FGDs of each country were trained on standard FGD methodology using a semi-structured FGD guide. The guide was based on the precaution adoption process model and the theory of planned behaviour,^11,12^ and consisted of questions exploring the concept of CS and existing CS activities related to pandemic management (Supplementary 1). Questions were also devised to understand the facilitators and barriers to the implementation of CS activities, as well as sustainability concerns. Participant information sheet, informed consent form, and FGD questions were translated from English to local languages (Nepali, Bangla, Hindi, Bahasa Indonesia, and Filipino) by the country team. Venues to conduct FGDs were selected by project staff to ensure convenience, privacy, and accessibility for study participants. Before the conduct of FGDs, participants were shown a video and infographics in the local languages explaining the meaning of CS, along with its various approaches, to acquaint the FGD participants with CS before responding to questions (Supplementary 2). A local country team comprising site investigators and project staff reviewed the study materials in local languages for clarity and understandability.

### Data analysis

Audio recordings of the FGDs were transcribed verbatim and translated from local languages to English. Collected data was coded and analyzed using qualitative content analysis methods through the steps of reading, coding, analysis, data display, and reduction as well as data interpretation, by trained project staff, supervised by the site investigators. The initial level of coding was done by the team to observe, compare, and identify similarities and differences in the data. Subsequently, second level of pattern coding was done by the country team for content analysis for participant’s narrative statement to extract subthemes and themes (main topic/subject/message). The initial findings were shared with other country teams in virtual workshops to validate analysis and coding. Finally, triangulation of patterns was done wherein subthemes and themes were refined and grouped based on similar patterns and meaning, after obtaining consensus from country teams to ensure face and content validity. It was done till thematic saturation was obtained i.e., no further refinement for better clarity was possible by country teams during workshops.

### Ethics Statement

Prior ethical approval was sought first from the Ethical Review Board (ERB), United Nations University Institute, Macau (Reference No: 202206/01), and subsequently from ethical/regulatory committee of each country. All participants gave informed consent and participated in focus group discussion. They agreed upon location and time of group discussion along with permission to audio record. Country-specific data and personal information are securely stored with the country team and no personal information of participants was shared while carrying out the analysis.

## Results

A total of 130 individuals participated in 20 FGDs across 5 countries (India: 27; Nepal: 25; Bangladesh: 29; Indonesia: 24; and Philippines: 25). On average, each FGD lasted for about 45 minutes (minimum: 35 minutes; maximum 90 minutes). While responding to FGD questions, participants related them with their perceptions for COVID-19 pandemic and subsequent response measures by their respective governments. Domains of FGD are summarized by the themes, subthemes, and illustrative quotes from the participants.

Firstly, concepts of CS and participants’ experiences and potential roles in pandemic preparedness and response were explored (Table 1):

**Table 1:**
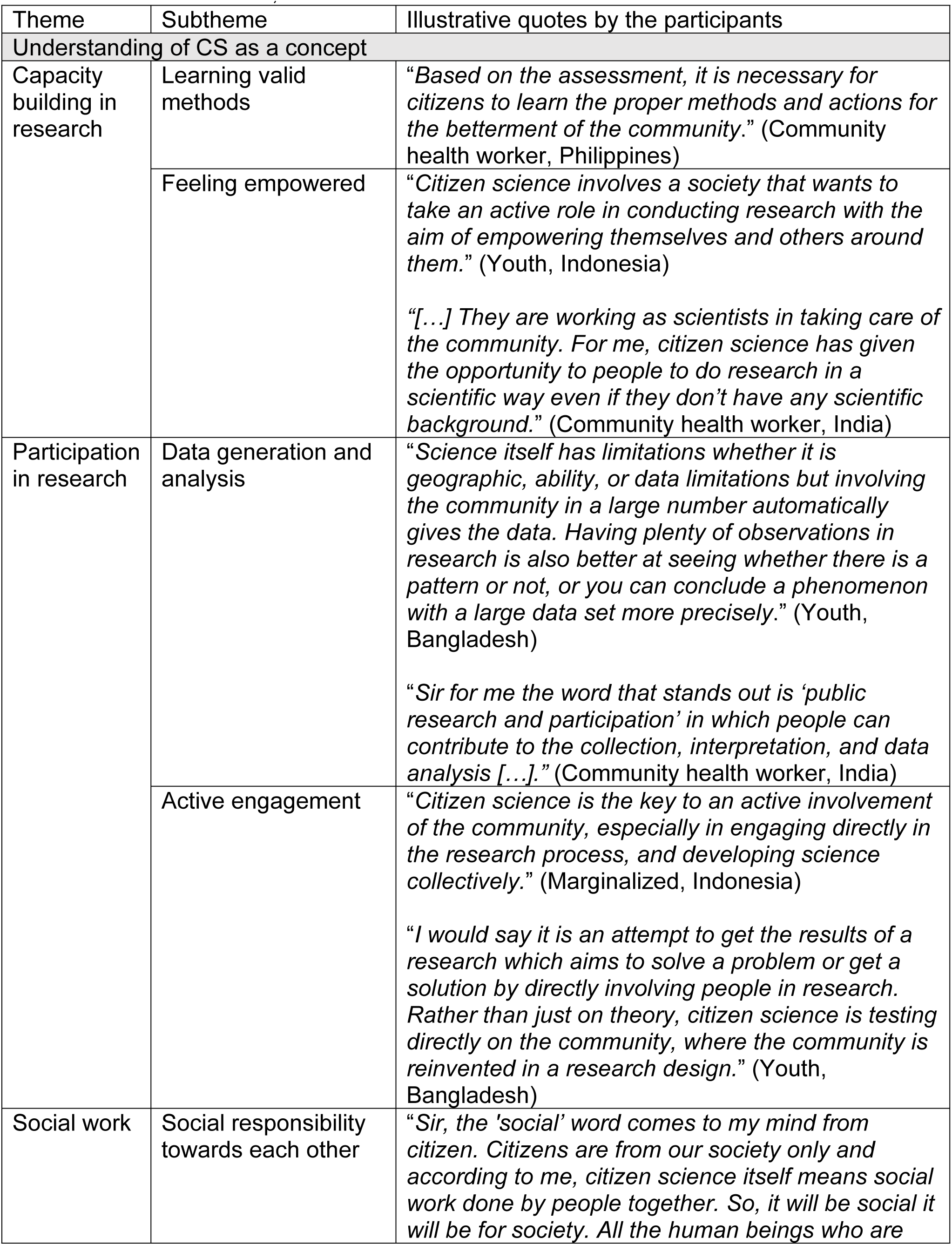
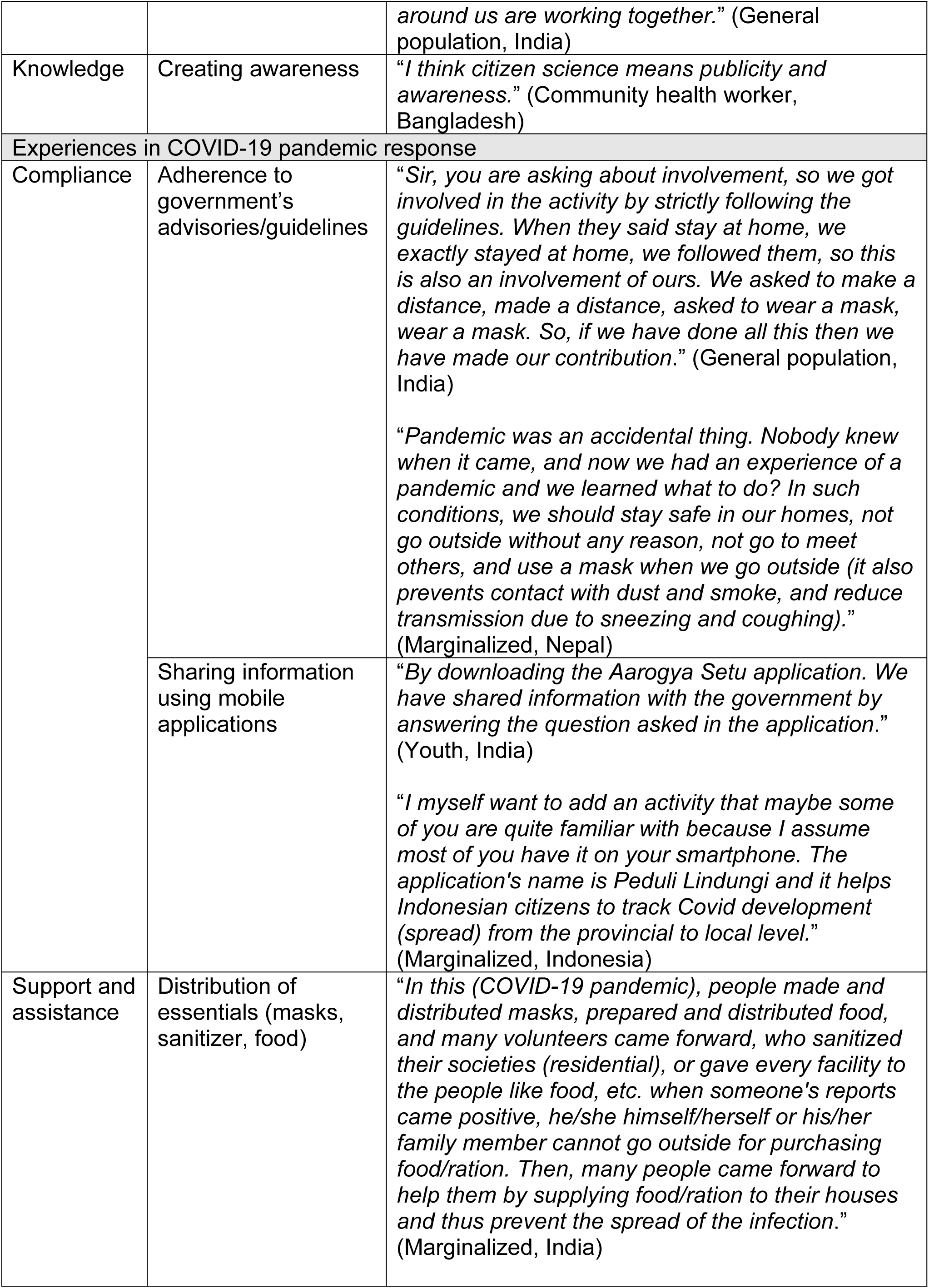
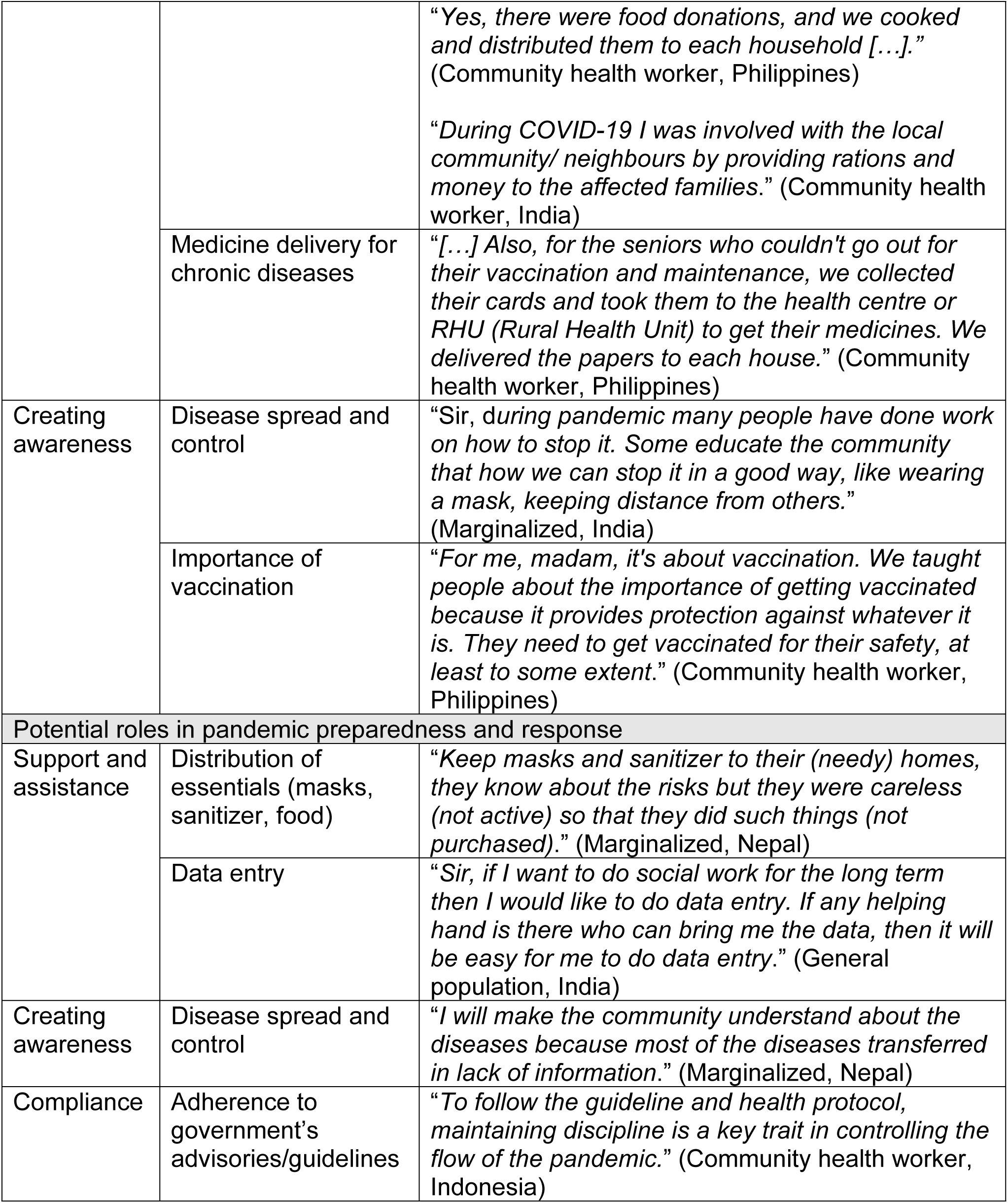
Thematic analysis of citizen science (CS) concept and participants’ potential roles in CS along with lived experiences during COVID-19 pandemic in South and Southeast Asian countries, 2022-23.

| Theme | Subtheme | Illustrative quotes by the participants |
| --- | --- | --- |
| Understanding of CS as a concept |  |  |
| Capacity building in research | Learning valid methods | *"Based on the assessment, it is necessary for citizens to learn the proper methods and actions for the betterment of the community."* (Community health worker, Philippines) |
|  | Feeling empowered | *"Citizen science involves a society that wants to take an active role in conducting research with the aim of empowering themselves and others around them."* (Youth, Indonesia)<br>*"[...] They are working as scientists in taking care of the community. For me, citizen science has given the opportunity to people to do research in a scientific way even if they don't have any scientific background."* (Community health worker, India) |
| Participation in research | Data generation and analysis | *"Science itself has limitations whether it is geographic, ability, or data limitations but involving the community in a large number automatically gives the data. Having plenty of observations in research is also better at seeing whether there is a pattern or not, or you can conclude a phenomenon with a large data set more precisely."* (Youth, Bangladesh)<br>*"Sir for me the word that stands out is 'public research and participation' in which people can contribute to the collection, interpretation, and data analysis [...]."* (Community health worker, India) |
|  | Active engagement | *"Citizen science is the key to an active involvement of the community, especially in engaging directly in the research process, and developing science collectively."* (Marginalized, Indonesia)<br>*"I would say it is an attempt to get the results of a research which aims to solve a problem or get a solution by directly involving people in research. Rather than just on theory, citizen science is testing directly on the community, where the community is reinvented in a research design."* (Youth, Bangladesh) |
| Social work | Social responsibility towards each other | *"Sir, the 'social' word comes to my mind from citizen. Citizens are from our society only and according to me, citizen science itself means social work done by people together. So, it will be social it will be for society. All the human beings who are* |
|  |  | *around us are working together.” (General population, India)* |
| Knowledge | Creating awareness | *“I think citizen science means publicity and awareness.” (Community health worker, Bangladesh)* |
| Experiences in COVID-19 pandemic response |  |  |
| Compliance | Adherence to government’s advisories/guidelines | *“Sir, you are asking about involvement, so we got involved in the activity by strictly following the guidelines. When they said stay at home, we exactly stayed at home, we followed them, so this is also an involvement of ours. We asked to make a distance, made a distance, asked to wear a mask, wear a mask. So, if we have done all this then we have made our contribution.” (General population, India)*<br>*“Pandemic was an accidental thing. Nobody knew when it came, and now we had an experience of a pandemic and we learned what to do? In such conditions, we should stay safe in our homes, not go outside without any reason, not go to meet others, and use a mask when we go outside (it also prevents contact with dust and smoke, and reduce transmission due to sneezing and coughing).” (Marginalized, Nepal)* |
|  | Sharing information using mobile applications | *“By downloading the Aarogya Setu application. We have shared information with the government by answering the question asked in the application.” (Youth, India)*<br>*“I myself want to add an activity that maybe some of you are quite familiar with because I assume most of you have it on your smartphone. The application's name is Peduli Lindungi and it helps Indonesian citizens to track Covid development (spread) from the provincial to local level.” (Marginalized, Indonesia)* |
| Support and assistance | Distribution of essentials (masks, sanitizer, food) | *“In this (COVID-19 pandemic), people made and distributed masks, prepared and distributed food, and many volunteers came forward, who sanitized their societies (residential), or gave every facility to the people like food, etc. when someone's reports came positive, he/she himself/herself or his/her family member cannot go outside for purchasing food/ration. Then, many people came forward to help them by supplying food/ration to their houses and thus prevent the spread of the infection.” (Marginalized, India)* |
|  |  | *"Yes, there were food donations, and we cooked and distributed them to each household [...]."* (Community health worker, Philippines)<br>*"During COVID-19 I was involved with the local community/ neighbours by providing rations and money to the affected families."* (Community health worker, India) |
|  | Medicine delivery for chronic diseases | *"[...] Also, for the seniors who couldn't go out for their vaccination and maintenance, we collected their cards and took them to the health centre or RHU (Rural Health Unit) to get their medicines. We delivered the papers to each house."* (Community health worker, Philippines) |
| Creating awareness | Disease spread and control | *"Sir, during pandemic many people have done work on how to stop it. Some educate the community that how we can stop it in a good way, like wearing a mask, keeping distance from others."* (Marginalized, India) |
|  | Importance of vaccination | *"For me, madam, it's about vaccination. We taught people about the importance of getting vaccinated because it provides protection against whatever it is. They need to get vaccinated for their safety, at least to some extent."* (Community health worker, Philippines) |
| Potential roles in pandemic preparedness and response |  |  |
| Support and assistance | Distribution of essentials (masks, sanitizer, food) | *"Keep masks and sanitizer to their (needy) homes, they know about the risks but they were careless (not active) so that they did such things (not purchased)."* (Marginalized, Nepal) |
|  | Data entry | *"Sir, if I want to do social work for the long term then I would like to do data entry. If any helping hand is there who can bring me the data, then it will be easy for me to do data entry."* (General population, India) |
| Creating awareness | Disease spread and control | *"I will make the community understand about the diseases because most of the diseases transferred in lack of information."* (Marginalized, Nepal) |
| Compliance | Adherence to government's advisories/guidelines | *"To follow the guideline and health protocol, maintaining discipline is a key trait in controlling the flow of the pandemic."* (Community health worker, Indonesia) |

a. Understanding of CS as a concept Across all countries, while referring to CS, the participants collectively comprehended the term "research". Participants understood that CS is possibly related to capacity building to learn and empower themselves in the field of research. CS as an engagement process was considered as an opportunity to participate in generating and analyzing data for effective implementation of control measures. It was also viewed as a social responsibility, especially in India. Distinctively, it was referred to as an awareness mechanism to improve knowledge about pandemic response in Bangladesh.
b. Experiences in COVID-19 pandemic response When participants were asked about their lived experiences during the pandemic in terms of nature of their engagement in pandemic response, majority of the participants (especially in India and Philippines) stated their participation by adhering to government guidelines in terms of following COVID-19 appropriate behaviour like wearing masks, maintaining physical distancing, and hand sanitization. They also responded that they complied with digital interventions especially downloading mobile-based applications to give personal information to assist the governments in tracing the spread of infection. Largely in India, participants said that they actively came forward to help fellow citizens and gave them support in terms of providing food, medicine, masks, and sanitizers. They shared about being part of a team with the local health authorities in organizing community-based awareness activities to provide knowledge about disease, its control measures, and importance of vaccination to reduce disease transmission.
c. Potential roles in pandemic preparedness and response After reflecting on their experiences, participants were asked about their likely roles in future pandemics. Across all countries, based on their lived experiences, most considered their likely participation to provide/assist food, masks, and sanitizers along with creating awareness. Participants also stated that as citizens, their role can be to ensure compliance by fellow citizens to government guidelines. Some stated that they can contribute based on their skill sets (e.g., computer skills for data entry) to assist pandemic preparedness and response.

After showing concept and meaning of CS using infographics, participants mentioned advantages and disadvantages of CS and highlighted potential facilitators and barriers to their participation in CS (Table 2):

**Table 2:** Thematic analysis of perceived issues (advantages, disadvantages, facilitators, and barriers) of citizen science (CS) to foster public partnerships for pandemic preparedness and response in South and Southeast Asian countries, 2022-23.

| Theme | Subtheme | Illustrative quotes by the participants |
| --- | --- | --- |
| **Advantages of CS** |  |  |
| People-centric | Realistic approach | *"They can gather more data or the pulse of the people. Because if you are just on top, you won't see what's happening below. But if you involve the people, you will know the actual situation in the community. You can get a better feel for the pulse of the people."* (Marginalized, Philippines)<br>*"Particularly people are familiarized with challenges and aspects that are affecting their daily lives and matters relating to their own intentions. How activities can be tiered to data and processed to make a policy."* (Youth, Indonesia) |
|  | Sharing of views | *"Sir, like-minded people group will be formed, people will come forward and everyone's views will be listened to."* (Youth, India) |
|  | Identifying solutions | *"Because the answers of the public can lead to a solution. So it's really necessary if we are somehow connected."* (Youth, Bangladesh) |
| Capacity Building | Gaining knowledge | *"The advantage of involving them is that they (people) gain knowledge about health and the right steps to take for their well-being. They learn what is best for them."* (Community health worker, Philippines)<br>*"It is necessary to involve the public in these activities to inform because most of the people didn't know the information about the pandemic."* (Marginalized, Nepal) |
|  | Creating awareness | *"Community awareness is the advantage of involving the public in research."* (General population, Philippines) |
| **Disadvantages of CS** |  |  |
| Ineffective participation | Not sharing of opinions | *"People keep the solution of a problem with them only and not share it with anyone."* (Youth, India) |
|  | Lack of capacity | *"My concern is that science works a bit differently so far because only people that are skilful are doing it. And not all people will understand the results of the research, people don't know about it at all."* (General population, Indonesia) |
| Risk of infection | Non-adherence to guidelines | *"Since the virus has unknown health implications for different groups, it is advisable to not involve the public actively in the early stage."* (Community health worker, Indonesia) |
|  |  | *"Social distancing, because we cannot socialize, it is difficult to socialize with other people because there is a risk of getting infected, things like that."*<br>(Youth, Philippines) |
| **Facilitators for participation in CS activities** |  |  |
| Individual factors | Achievement | *"One of the factors is a sense of achievement that my voice is being listened to and implemented."*<br>(Youth, India) |
|  | Satisfaction | *"Emotional, feeling empowered in control of an issue when you join here because you feel like 'I joined here' like I was able to help the community, it is like you feed your ego and now you are satisfied."* (Youth, Philippines) |
|  | Happiness | *"As we are still young, at this age, I would like to help others, because we get eternal happiness in helping others. If someone praises us or if someone gets cured due to my help, then I will be very happy."* (General population, India) |
|  | Gain new knowledge | *"For me, the first main reason is that hopefully, this research can bring a change for us in the future. The second reason is that this is a recipe ingredient for researchers in terms of operational development which will be required later, and the last thing is that we can get knowledge too. In citizen science, usually, the results we can read for our reference and increase our knowledge too."*<br>(General population, Indonesia)<br>*"To be able to learn new knowledge and skills. To acquire new learning."* (Marginalized, Philippines)<br>*"I see the benefits, the second is to add insight and knowledge [...]"* (Community health worker, Indonesia) |
|  | Build social networks | *"[...] And the third is that we carry out social activities to increase relations and networking."*<br>(Community health worker, Indonesia) |
|  | Social duty and service | *"It is a social duty. I am an emotional person, so I use to participate in such kinds of activities. Popularity is minor for me."* (Youth, Nepal)<br>*"Main thing is social service. Human is social animal so he/she searches for social service. Other thing is, cultural, and attitudes are also related to social."* (General population, Nepal) |
| **Barriers to participation in CS activities** |  |  |
| Individual factors | Lack of education | *"Lack of education which creates trouble to make them understand."* (Marginalized, Nepal) |
|  | Lack of Information | *"The main reason for not participating may be because of the lack of information." (Youth, Indonesia)* |
|  | Lack of confidence | *"Lack of initiative. A person itself does not want to help anyone or put his/her personal opinion in front of anyone." (Youth, India)* |
|  | Lack of time | *"As we are working women, we have to come to the office from 10 AM to 5 PM, before that we have to do household work, look after the children, then when we reach home in the evening, we have to prepare food, taught children, etc. So, we do not get time to participate in such activities, it pushes us back." (Marginalized, India)*<br>*"Yes, if I am, maybe the most important reason is that the first is a matter of time. For people who work (job), there are time-related problems, scheduling problems, and how long for example we will have to participate. Don't mind doing it during free time outside working hours." (General population, Indonesia)* |
|  | Demotivation | *"If I get motivated by something then also get discouraged by that thing, like if I do some social work, many people are helping in that and many do not. Those who do not help, then I feel discouraged that if they are not helping, then why should I do the same?" (General population, India)* |
|  | Risk of infection | *"Sir, I wanted to help during Covid, but I had a fear that I might get infected. If I had it, then the small children in my house may also get infected. So, I did not participate because I was afraid that this thing might happen to me and my family." (Marginalized, India)* |
| Organizational factors | Unengaging organizers | *"Because there are organizers who are boring when teaching. And sometimes I do not know if they are just lazy to teach or if they don't want to teach what they know. The same goes for me, I prefer someone who is lively because being a killjoy is not allowed here." (Marginalized, Philippines)* |
|  | Lack of transportation | *"For me, an example would be when the travel distance is far, and there is no immediate transportation available to get off, but there must be a way, but that is the most common reason for me, the distance, the location." (Community health worker, Philippines)* |

a. Advantages and disadvantages of CS While speaking about advantages, across all countries, participants stated that CS is a people-centric approach that can be used to understand reality on the ground, gather people to encourage exchange opinions, and identify people-led solutions. Potentially, CS can build the capacity of people by generating knowledge and creating awareness about their health and pandemic control measures. The disadvantages of CS were largely linked to individual factors such as lack of capacity or unwillingness to share opinions that will likely cause ineffective participation. Based on their COVID-19 experience, participants expressed a potential risk of infection to themselves and their families while gathering people in CS activities.
b. Facilitators and barriers to participation in CS activities While assessing for potential facilitating factors, participants again expressed individual-related factors. In India, CS activities are expected to instil a sense of achievement, satisfaction, and happiness. In Philippines, potential chances to gain new knowledge and build social networks were also considered to be facilitators. Whereas, in India and Nepal, majority of the participants stated that a sense of social duty and service during a pandemic enables participation. Similar to facilitators, individual-related factors were also considered to be barriers to CS activities. Nature of concerns varied across countries, such as the lack of information in Bangladesh, Indonesia, and Philippines; lack of education, confidence, and individual level conflicts in India and Nepal; while lack of time to participate was expressed as a barrier across all countries. Participants also mentioned that the nature of facilitation can have an impact on the level of participation. In Nepal, participants said that they tend to feel demotivated if other participants choose not to participate, while one is making an effort. In addition to individual-related factors, organizational factors such as unengaged organizers/facilitators and lack of transportation assistance can pose barriers to participate.

Finally, factors related to interactions with stakeholders (Table 3) and sustainability and resources needed (Table 4) to foster strong public partnerships for pandemic preparedness and response were discussed:

**Table 3:**
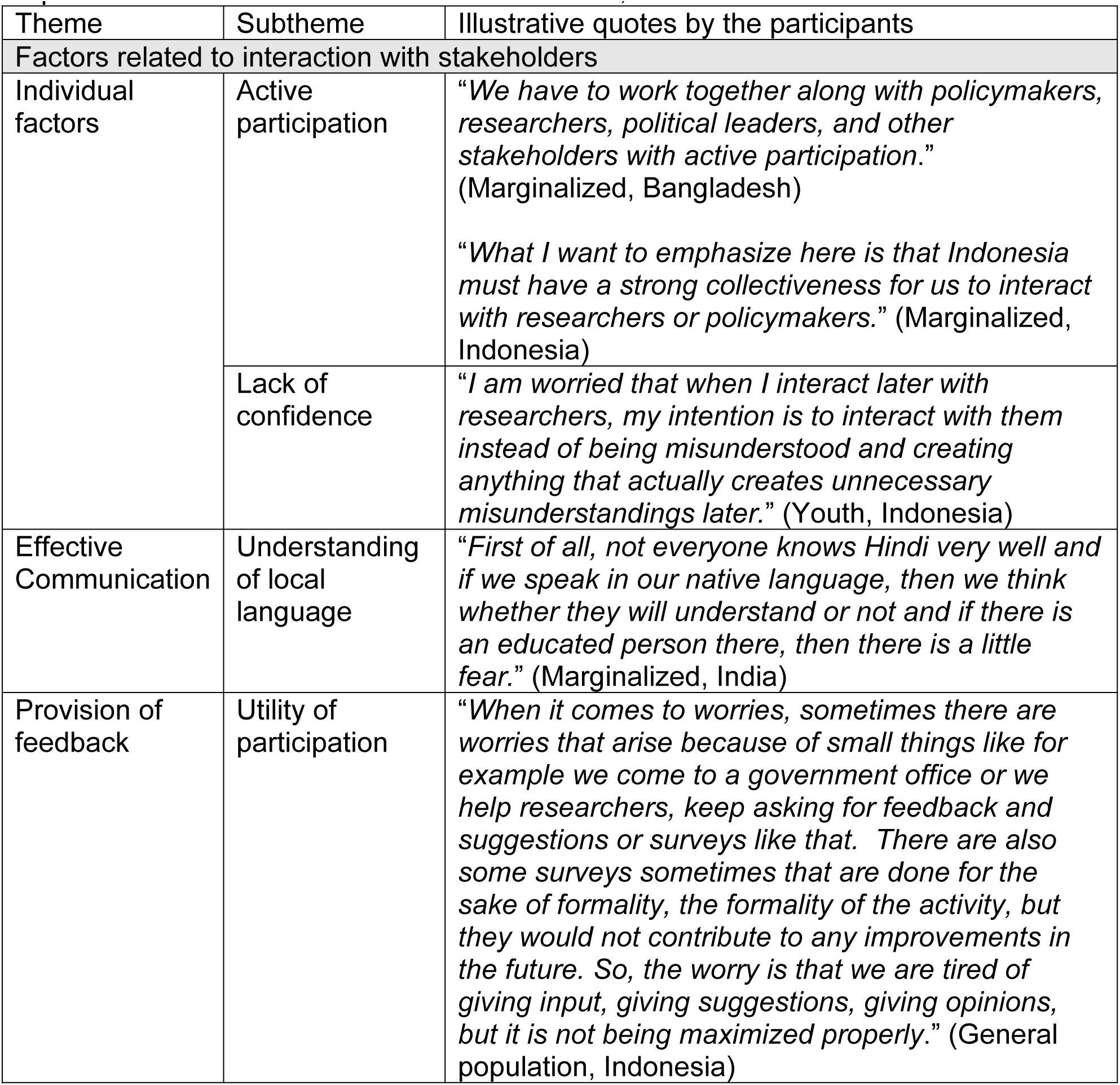
Thematic analysis of factors related to interaction with stakeholders for citizen science (CS) to foster public partnerships for pandemic preparedness and response in South and Southeast Asian countries, 2022-23.

| Theme | Subtheme | Illustrative quotes by the participants |
| --- | --- | --- |
| Factors related to interaction with stakeholders |  |  |
| Individual factors | Active participation | *"We have to work together along with policymakers, researchers, political leaders, and other stakeholders with active participation."* (Marginalized, Bangladesh)<br>*"What I want to emphasize here is that Indonesia must have a strong collectiveness for us to interact with researchers or policymakers."* (Marginalized, Indonesia) |
|  | Lack of confidence | *"I am worried that when I interact later with researchers, my intention is to interact with them instead of being misunderstood and creating anything that actually creates unnecessary misunderstandings later."* (Youth, Indonesia) |
| Effective Communication | Understanding of local language | *"First of all, not everyone knows Hindi very well and if we speak in our native language, then we think whether they will understand or not and if there is an educated person there, then there is a little fear."* (Marginalized, India) |
| Provision of feedback | Utility of participation | *"When it comes to worries, sometimes there are worries that arise because of small things like for example we come to a government office or we help researchers, keep asking for feedback and suggestions or surveys like that. There are also some surveys sometimes that are done for the sake of formality, the formality of the activity, but they would not contribute to any improvements in the future. So, the worry is that we are tired of giving input, giving suggestions, giving opinions, but it is not being maximized properly."* (General population, Indonesia) |

**Table 4:**
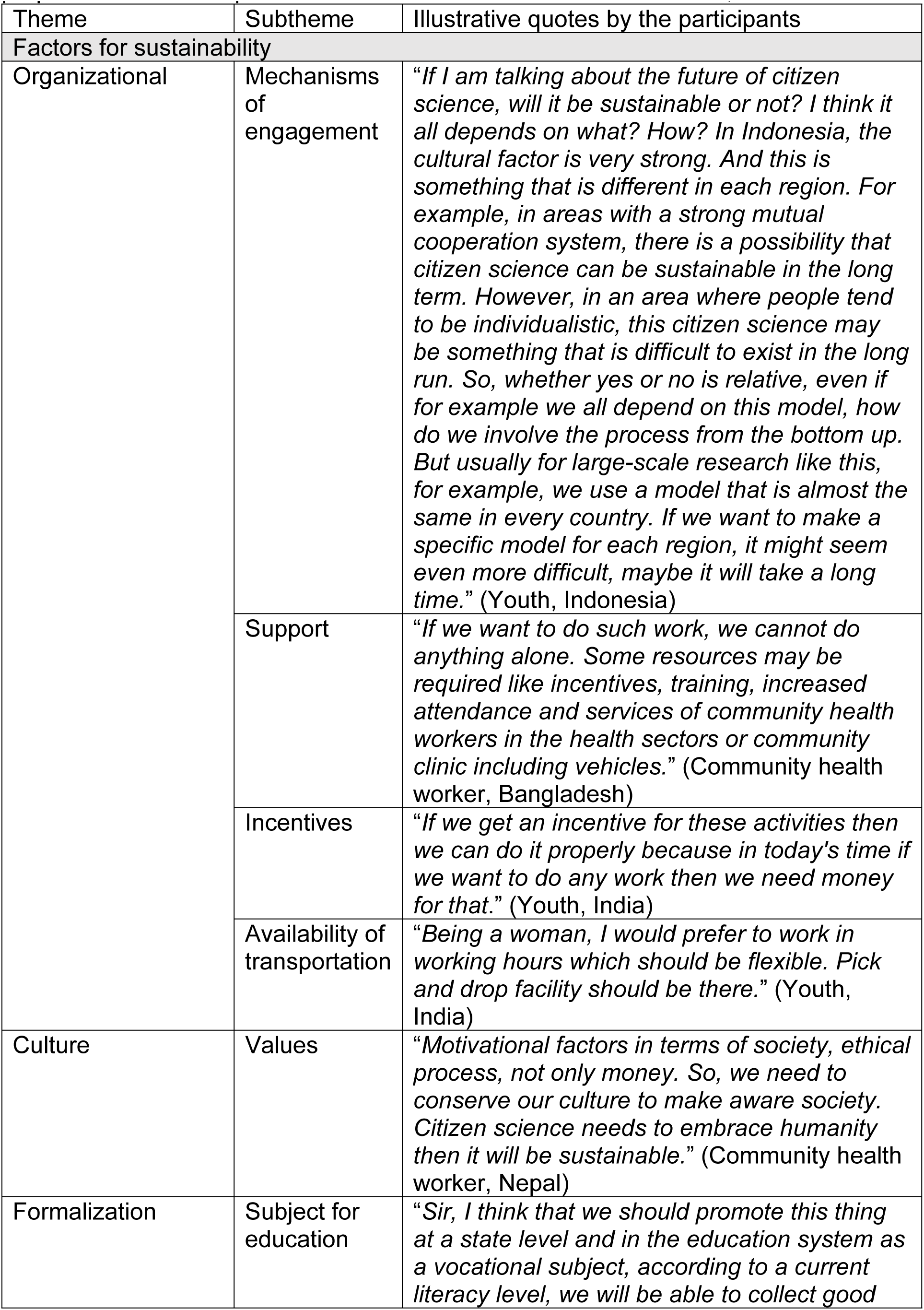
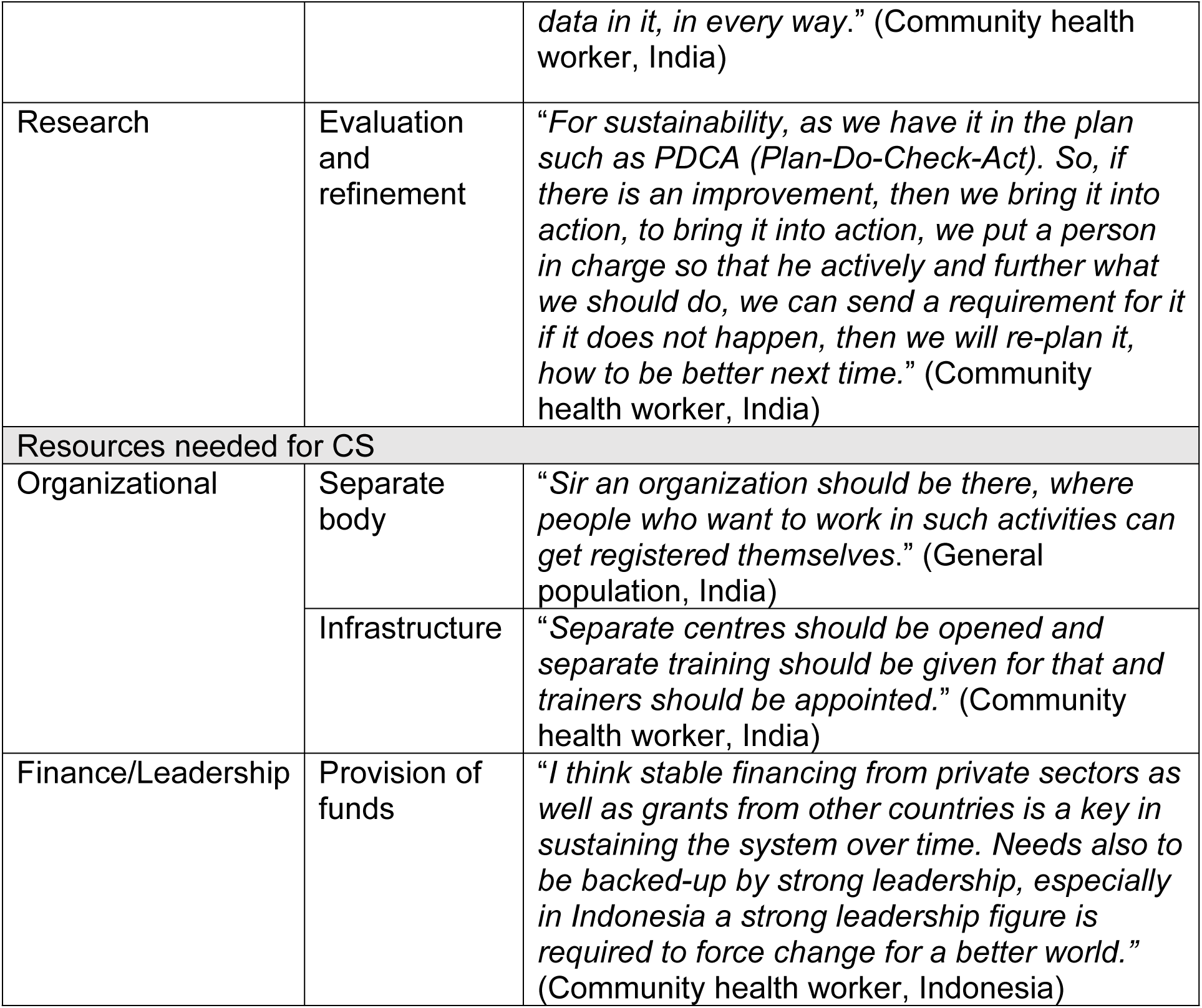
Thematic analysis of perceived sustainability factors of and needed resources for citizen science (CS) to foster public partnerships for pandemic preparedness and response in South and Southeast Asian countries, 2022-23.

| Theme | Subtheme | Illustrative quotes by the participants |
| --- | --- | --- |
| **Factors for sustainability** |  |  |
| Organizational | Mechanisms of engagement | *"If I am talking about the future of citizen science, will it be sustainable or not? I think it all depends on what? How? In Indonesia, the cultural factor is very strong. And this is something that is different in each region. For example, in areas with a strong mutual cooperation system, there is a possibility that citizen science can be sustainable in the long term. However, in an area where people tend to be individualistic, this citizen science may be something that is difficult to exist in the long run. So, whether yes or no is relative, even if for example we all depend on this model, how do we involve the process from the bottom up. But usually for large-scale research like this, for example, we use a model that is almost the same in every country. If we want to make a specific model for each region, it might seem even more difficult, maybe it will take a long time."* (Youth, Indonesia) |
|  | Support | *"If we want to do such work, we cannot do anything alone. Some resources may be required like incentives, training, increased attendance and services of community health workers in the health sectors or community clinic including vehicles."* (Community health worker, Bangladesh) |
|  | Incentives | *"If we get an incentive for these activities then we can do it properly because in today's time if we want to do any work then we need money for that."* (Youth, India) |
|  | Availability of transportation | *"Being a woman, I would prefer to work in working hours which should be flexible. Pick and drop facility should be there."* (Youth, India) |
| Culture | Values | *"Motivational factors in terms of society, ethical process, not only money. So, we need to conserve our culture to make aware society. Citizen science needs to embrace humanity then it will be sustainable."* (Community health worker, Nepal) |
| Formalization | Subject for education | *"Sir, I think that we should promote this thing at a state level and in the education system as a vocational subject, according to a current literacy level, we will be able to collect good* |
|  |  | *data in it, in every way.” (Community health worker, India)* |
| Research | Evaluation and refinement | *“For sustainability, as we have it in the plan such as PDCA (Plan-Do-Check-Act). So, if there is an improvement, then we bring it into action, to bring it into action, we put a person in charge so that he actively and further what we should do, we can send a requirement for it if it does not happen, then we will re-plan it, how to be better next time.” (Community health worker, India)* |
| Resources needed for CS |  |  |
| Organizational | Separate body | *“Sir an organization should be there, where people who want to work in such activities can get registered themselves.” (General population, India)* |
|  | Infrastructure | *“Separate centres should be opened and separate training should be given for that and trainers should be appointed.” (Community health worker, India)* |
| Finance/Leadership | Provision of funds | *“I think stable financing from private sectors as well as grants from other countries is a key in sustaining the system over time. Needs also to be backed-up by strong leadership, especially in Indonesia a strong leadership figure is required to force change for a better world.” (Community health worker, Indonesia)* |

a. Factors related to interaction with stakeholders When asked about factors to consider when interacting with stakeholders such as policymakers and researchers, most factors were related to individual characteristics, effective communication, and provision of feedback. Active participation and a strong sense of collectiveness were raised as important factors. In India and Indonesia, contextual awareness of stakeholders and their ability to correctly understand participants were shared as likely factors. Participants expressed their lack of confidence while sharing opinions with stakeholders due to differences in their level of education and language which is likely to result in misunderstandings. In Indonesia, participants expressed concerns about lack of utilization of their feedbacks even though they had previously shared their opinions, data, and information in various activities like surveys.
b. Sustainability factors and resources needed for CS When asked to express potential sustainability factors, the importance of local mechanisms to sustain people’s participation in CS activities was mentioned in Indonesia. The need for health system support, provision of incentives (monetary and non-monetary), and transportation assistance, especially to women, were delineated for CS sustenance in India and Bangladesh. Contextually, in India and Nepal, cultural factors were expected to play a vital role in the sustainability of CS. In India, introduction of CS in formal education system as a distinct identity was also considered as a sustainability factor to teach concepts and applications of CS. A participant from India suggested that the use of a scientific approach to monitor and evaluate CS activities for concurrent refinements can further enhance sustainability. The requirement of resources was substantiated to initiate and sustain CS activities. A formal structure such as the establishment of an organization or network and creation of infrastructures and centres along with budgetary support were considered as resources needed for CS in pandemic preparedness and response.

## Discussion

Current study captured COVID-19 lived experiences and qualitatively explored the potential application of CS approach to encourage people’s participation in pandemic preparedness and response. The understanding of CS, issues related to citizens’ participation, resources needed, and sustainability factors were discussed with the study participants. Contextual sensitiveness was considered in the present study with the inclusion of South and Southeast Asian countries. Population representativeness was ensured by including diverse groups such as youth, general population, marginalized and indigenous groups, and community health workers. To the best of our knowledge, there has been a lack of examination of the citizens’ perspectives toward the CS approach in pandemic preparedness and response. Hence, this study contributes to the knowledge gap in applications of CS in the pandemic context.

Thematic analysis of FGDs involving participants across five countries gave an insight into the various facets of CS and its applications. CS was expressed as a social concept, especially in India and Nepal. Across all countries, CS was viewed to have potential to capacitate and empower citizens while working with researchers. It was largely appreciated in the backdrop of their lived experiences during the COVID-19 pandemic. Mostly in India and Nepal, their participation in its control was not only limited to comply with government guidelines but was extended to assist people with masks, sanitizers, food, and medicine. During pandemic, people created awareness among citizens and shared personal information about their COVID-19 status to assist governments in monitoring the disease spread. These experiences shaped their responses toward CS as COVID-19 resonated in their voices and quotes. CS was considered to be a people-centric approach with the advantage of helping the policymakers reduce disease transmission with citizens’ participation. Across all countries, individual-level factors such as a sense of satisfaction, social duty, and potential to build their capacity were considered facilitating factors to participate in CS activities. Major disadvantages and barriers to partake in activities were lack of time, confidence, and capacity (knowledge/skills), insufficient transportation assistance, and risk of infection.

Individual-level factors that are related to the interaction and sharing of opinions with stakeholders such as policymakers and researchers include active participation and attitude of participants, communication and fear of misunderstanding, and actual application of their responses in implementing strategies. Across all countries, provision of incentives and transportation assistance were reflected to sustain CS activities, along with resources like organizational support and funding. In Indonesia, participants expressed that CS activities will be sustainable when they take into account local contexts and are developed based on existing mechanisms.

Findings from this study resonated with the literature. Individual-level factors like self-esteem and self-efficacy along with health status were observed to be related to community participation.^13^ Level of participation was found to be affected by availability, geographic location, and socio-economic status of community.^14^ Subjective norms, cognitive ability of participants, experience of disaster, and perception of risk were delineated as individual-level factors for participation.^15^ Community engagement in emergencies was observed to be improved with accessibility to internet. It showed further improvement in sustenance of availability of health care services.^16^ Evidence also showed that enhanced community participation is affected by an established management system, capacity building of community, and experiences and vulnerability of disaster.^17^ Current study summates that COVID-19 experiences gave rationale and ways to raise resources to conduct CS activities for meaningful and effective participation.

CS makes public participation in science more democratized while involving researchers makes it more rational and objective.^18^ People being the recipients of policies and programs should have their voices included. Readiness of citizens should be ensured by prior and repeated sensitization about CS approaches and ways of participation. To provide local resonance with the CS approach, it is vital to consider the contextual settings while studying the values, norms, and culture. Individual capacity in terms of baseline knowledge and skill, awareness, and cognition needs to be mapped to tailor engagement efforts. It is evident from current analysis that people are ready to participate but they need to be informed and their fears (infection, miscommunication, and being misunderstood by stakeholders) needs to be addressed. Efforts need to be targeted to make people comfortable and respected without any language barriers. Measures need to be in place to identify/foresee potential conflicts between people and train for conflict resolution strategies to make participation efficient and effective. A formal feedback structure should also be available to inform the public about the utility of their participation and the inclusion of their opinions in formulating interventions and policies. It is evident that feedback increases the level of motivation and it should be specific with reasons or criterion-based.^19^

Current study has its strengths in terms of in-depth examination with a structured inquiry through conduct of FGDs by local teams in local languages. Participants were contacted after obtaining consent while carrying out the quantitative survey. Study had a sufficient sample size with enriched narrative quotes representative of the population (youth, marginalized and indigenous, and general) along with community health workers. It covered provider and beneficiary perspectives and experiences during the COVID-19 pandemic, and gave relatable responses towards pandemic preparedness and control. Similar and differing views of CS were analyzed and compared across five countries. Although generalization of qualitative information poses one of the limitations along with management of voluminous data.^20^ Our findings can be triangulated with the quantitative segment of our mixed-method study.^10^ Country-specific teams led by site researchers maintained rigor for data collection, and thematic analysis of quotes made analysis systematic and contextual. The concept of CS was relatively new to most participants making it hard for some to understand and relate to the questions. We tried to mitigate this by introducing CS at the start with a video and infographics in local languages.

## Conclusions

In-depth analysis suggested that people are ready to participate with stakeholders such as policymakers and researchers, considering their social duty driven by attitude and culture. Based on their experiences during COVID-19, participation in CS activities viewed as an opportunity to learn new knowledge and scientific skills while working with researchers. Discomfort while communicating with other stakeholders was conveyed due to limited knowledge and skills of participants. The need for CS organisation or network, inclusion in education system, funding provisions, and incentives to participants were shared as sustainable factors and resources. As expected, participants’ responses in current study resonated with the COVID-19 pandemic. Future work can explore the application of CS in other disease outbreaks/epidemics.

## Data Availability

Data can be provided upon request.

## Acknowledgements

Authors would like to thank people who participated in study along with members of community-based organizations.

## Author contributions

All authors contributed to the conception of this study and the development of all study materials. YRT and PY oversaw the study and had full access to all data. DK, IH, FCC, FYS, GRM, MMA, and HK were responsible for data collection in their respective countries and had access to their own country’s data. All authors contributed to data analyses. DK drafted the paper, which was commented upon by all authors prior to submission. All authors gave the final approval of the version to be published.

